# Point-of-care lung ultrasound predicts severe disease and death due to COVID-19: a prospective cohort study

**DOI:** 10.1101/2021.12.30.21268558

**Authors:** Paul W. Blair, Trishul Siddharthan, Gigi Liu, Jiawei Bai, Joshua East, Phabiola Herrera, Lalaine Anova, Varun Mahadevan, Shakir Hossen, Stefanie Seo, Olamide Sonuga, Joshua Lawrence, Jillian Peters, Andrea Cox, Yukari C. Manabe, Katherine Fenstermacher, Sophia Shea, Richard E. Rothman, Bhakti Hansoti, Lauren Sauer, Ciprian Crainiceanu, Danielle V. Clark

**Affiliations:** The Henry M. Jackson Foundation for the Advancement of Military Medicine, Bethesda, MD; Division of Infectious Diseases, Johns Hopkins University School of Medicine, Baltimore, MD; Division of Pulmonary and Critical Care Medicine, University of Miami, Miami, FL; Division of Pulmonary and Critical Care Medicine, Johns Hopkins University School of Medicine, Baltimore, MD; Division of General Internal Medicine, Johns Hopkins University School of Medicine, Baltimore, MD; Department of Biostatistics, Johns Hopkins Bloomberg School of Public Health, Baltimore MD; Department of Emergency Medicine, Johns Hopkins University, Baltimore MD

**Keywords:** COVID-19, Ultrasonography, SARS-CoV-2, Survival Analysis, Cohort Studies

## Abstract

**Objective:** The clinical utility of point-of-care lung ultrasound (LUS) for disease severity triage of hospitalized patients with COVID-19 is unclear.

**Design:** Prospective cohort study

**Setting:** A large tertiary care center in Maryland, USA between April 2020 to September 2021.

**Patients:** Hospitalized adults (≥18 years of age) with positive SARS-CoV-2 RT-PCR results.

**Interventions:** None.

**Measurements and Main Results:** All patients were scanned using a standardized protocol including 12 lung zones and followed to determine clinical outcomes until hospital discharge and vital status at 28-days. Ultrasounds were independently reviewed for lung and pleural line artifacts and abnormalities, and the mean Lung Ultrasound Score (ranging from 0 to 3) across lung zones (mLUSS) was determined. The primary outcome was time to ICU-level care, defined as high flow oxygen, noninvasive, or mechanical ventilation, within 28-days of the initial ultrasound. Cox proportional hazards regression models adjusted for age and sex were fit for mLUSS and each ultrasound covariate. A total of 264 participants were enrolled in the study; the median age was 59 years and 114 (43.2) % of participants were female. The median mLUSS was 1 (interquartile range: 0.5 to 1.3). Following enrollment, 29 (11.0%) participants went on to require ICU-level care and 14 (5.3%) subsequently died by 28 days. Each increase in mLUSS at enrollment was associated with disease progression to ICU-level care (aHR = 3.63; 95% CI: 1.23 to 10.65) and 28-day mortality (aHR = 4.50; 95% CI: 1.52 to 13.31). Pleural line abnormalities were independently associated with disease progression to ICU-level care (aHR = 18.86; CI: 1.57 to 226.09).

**Conclusions:** Participants with a mLUSS ≥1 or pleural line changes on LUS had an increased likelihood of subsequent requirement of high flow oxygen or greater. LUS is a promising tool for assessing risk of COVID-19 progression at the bedside.

## Introduction

Point-of-care lung ultrasound (LUS) has been used for the evaluation of a range of cardiopulmonary conditions in emergency and critical care settings though, to date, implementation protocols have varied across settings. LUS offers benefits over traditional imaging modalities including portability, instantaneous results, lower costs, and lack of exposure to ionizing radiation. LUS has been proposed as an essential tool in evaluating patients with coronavirus disease 2019 (COVID-19) pneumonia to prevent nosocomial spread of disease.(1) Ultrasound hardware can be cleaned easily and reduces the burden on personnel and resources that would be required for traditional chest imaging. LUS may be able to identify patients at risk for decompensation requiring higher level of care in resource-limited settings or in regions with limited ICU capacity during a COVID-19 surge.

Despite the potential utility of LUS in COVID-19 management, standardized and evidence-based clinical use has not been fully established. The most widely studied and reported findings are based on the LUS score, originally developed in 2011 and used for assessment of aeration for titration of positive end-expiratory pressure (PEEP) (2). This scoring system includes a 0 to 3 point grade per 6 lung zones totaled from each hemithorax (3). This has been adopted for prognostication for non-COVID-19 acute respiratory distress syndrome (4) and was subsequently evaluated as a part of candidate models for COVID-19 prognostication (5-9). Among individuals with COVID-19, the LUS score has been associated with relevant chest CT findings and predicts the extent of parenchymal disease as well as mortality.(5) However, modified scores have limitations and have not been widely adopted. Modifications to scores had been based on early anecdotal reports and resulted in multiple scoring systems without protocol standardization and unclear generalizability. The LUS scores predicate on being able to sum all 12 zones, which can be challenging to obtain in tenuous patients in prone position.

The aim of this study was to determine the association between baseline lung ultrasound findings and the ultimate oxygen support requirements or death. We used a mean LUS (mLUSS) rather than a sum to determine the utility of the original and most studied LUS score (2) for COVID-19 prognostication with the added flexibility to include less than 12 lung zones. We performed a survival analysis with Cox regression mLUSS to determine risk of subsequently requiring ICU-level care (i.e., either high flow oxygen, noninvasive, or invasive ventilation) as a primary outcome. Secondary outcomes included ventilation plus 28-day death or 28-day death alone. We hypothesized that the mLUSS would be associated with an increased risk of progression to requiring ICU-level care.

## Methods

We enrolled adults (≥18 years of age) who tested positive for SARS-CoV-2 on RT-PCR and were admitted to Johns Hopkins Hospital in Baltimore, Maryland into a larger COVID-19 prospective cohort after verbal informed consent, between April 2020 to September 2021 as a convenience sample. This protocol was approved by the Johns Hopkins University Institutional Review Board (IRB00245545). Participants were enrolled after admission throughout the enrollment period or from the emergency department starting December 2020. After screening 2,270 patients, 723 participants enrolled into the master protocol, and 264 of these participants had LUS performed as part of study procedures depending on LUS-trained research staff availability.

LUS was standardized with 6-second clips from 12 lung zones with six lung zones on each side as previously described.(10) All images were collected with a Lumify S4 phased array probe (Philips, Amsterdam, Netherlands) using the application’s lung scan settings. We employed a standardized, point-of-care ultrasound (POCUS) research protocol to characterize lung abnormalities in COVID-19. Study personnel received training by a clinician certified in critical care ultrasonography and reviewed the initial ultrasound scanning sessions until operators were proficient. Study personnel were subsequently masked to clinical information and recorded LUS reads identifying and characterizing A lines (Figure 1A), B lines (Figure 1B and 1C), pleural effusions (Figure 1D), pleural line abnormalities (Figure 1E), and consolidations (Figure 1F). The pleural line was considered abnormal if it was irregular, fragmented, discontinuous, or ≥0.5 cm in thickness. Consolidations were required to be ≥0.25 cm in at least one dimension. While hospitalized, study visits including lung ultrasound scans occurred on study days 0, 3, 7, and weekly for up to 90 days. The first available scan was used for this analysis. Baseline demographics, comorbid conditions, and oxygen requirements until discharge were determined using the Hopkins Precision Medicine Analytics Platform (11), and duration of symptoms at enrollment was determined through medical chart review. Date of death by 28 days from enrollment was determined using the Precision Medicine Analytics Platform, medical chart review, and review of the regional Maryland, Washington D.C, and Virginia health information exchange (12).

**Figure 1.**
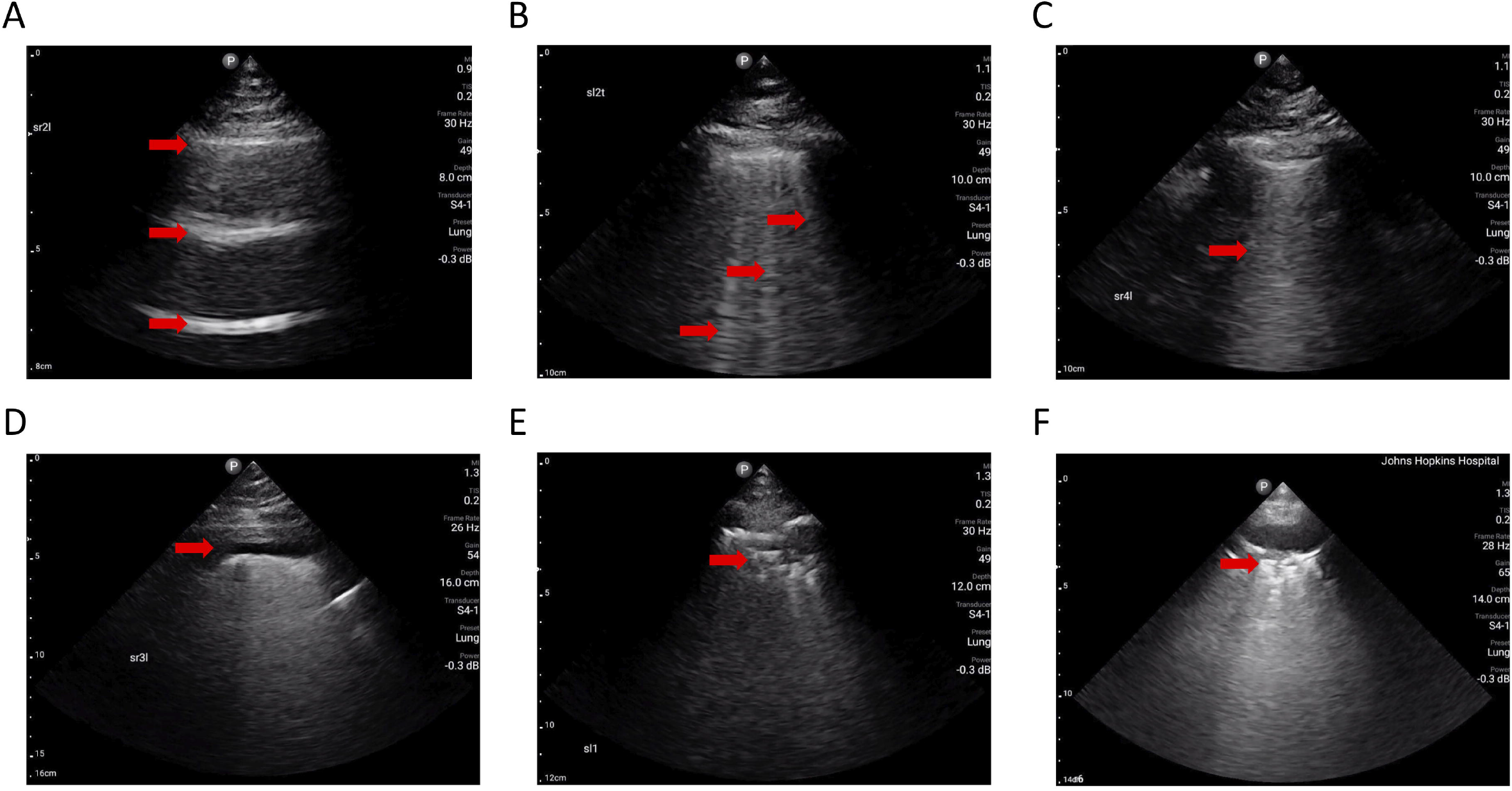
Lung ultrasound characteristics. 1A: A lines (arrows) are shown indicating normal lung aeration; 1B: discrete B lines (arrows), implicating subpleural interstitial edema; 1C: a coalesced B line (arrow); 1D: a small pleural effusion (arrow); 1E: fragmented pleural line (arrow); 1F: a subpleural consolidation (arrow).

As previously described (13), the LUS score was calculated for each zone with 1 point for discrete B lines, 2 points for coalescent B lines, and 3 points for lung consolidation. The mean LUS score (mLUSS) ranges from 0 to 3, with a higher score signifying higher severity. The mLUSS was calculated out of total available zones to include participants with missing zones. The Pearson correlation coefficient of the mLUSS between masked ultrasound clip readers was determined for the participants that were available (61 consecutive patients or 23% of the cohort). Participants were divided into severity groups at baseline based on severity at the time of POCUS or peak severity prior to POCUS: on room air or nasal cannula supplemental oxygen (moderate disease), on HFNC or noninvasive positive pressure ventilation (NIPPV) (moderately severe), or on mechanical ventilation (severe disease). Summary statistics were performed by comparing baseline demographics (i.e., sex, age, race, ethnicity, medical comorbidities), and duration post symptom-onset between severity groups using Kruskal-Wallis tests.

Progression to ICU-level care was defined as newly requiring either high flow nasal cannula, noninvasive ventilation, or mechanical ventilation during the hospitalization. To determine the association between baseline LUS characteristics and future risk, this outcome was restricted to study participants not requiring more than supplemental oxygen via low-flow nasal cannula at baseline (among those with moderate disease at baseline, N=164) (Figure 2). Secondary outcomes included 28-day mortality (all baseline severity groups, N=264) and 28-day progression to mechanical ventilation or 28-day death (among those with moderate or moderately severe disease, N=215) (Figure 2). A Kaplan-Meier plot was created to compare risk over time between those at the 25^th^ and 75^th^ mLUSS percentile. After checking the proportional hazards assumption, Cox proportional hazards regression models were used to evaluate the differences in risk of death and risk of death or subsequent mechanical ventilation plus 28-day death as a function of baseline % of lung fields with A lines, % with B lines, % with consolidations, % with pleural line abnormalities, % with pleural effusions, or the mLUSS. Unadjusted analyses and analyses adjusting for age and biologic sex were performed. A sensitivity analysis of the model including age, sex, and mLUSS covariates was performed restricting the population to those with all 12 lung zones for each of the primary and secondary outcomes. An additional sensitivity analysis was performed excluding 16 participants that were asymptomatic. Data were analyzed in R (v4.0.2) and Stata, version 16.0 (StataCorp LLC, College Station, TX, USA).

**Figure 2.**
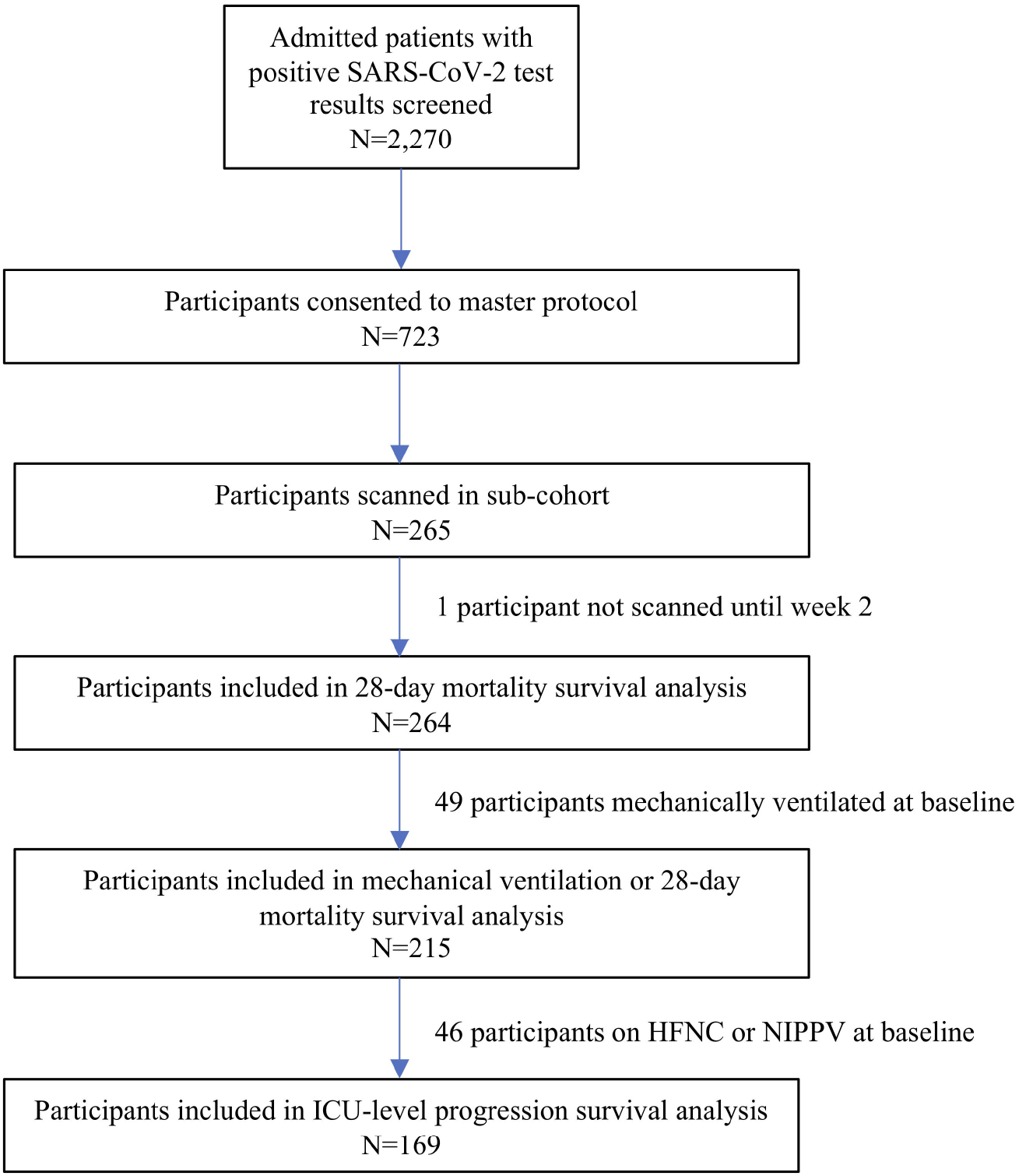
Flow diagram of enrollments and total participants included in survival analyses.

## Results

Of 264 participants, the median age was 61 (interquartile range [IQR], 48 to 68 years) and 43.0% (n = 114) were female (Table 1). The study participants were racially and ethnically diverse with 47.9% (n=127) of the population identified as black and 16.6% (n = 44) identified as Hispanic. The median time from symptoms onset until ultrasound scan was 9.29 days (IQR, 5.15 to 14.31 days) and the median mLUSS at baseline was 1.00 (IQR, 0.50 – 1.30) overall. Comorbid illness was common. The majority (74.2%) of participants had hypertension and 42.4% participants had diabetes mellitus (Table 1). Diagnoses of congestive heart failure (33.0%) and chronic obstructive pulmonary disease (36.4%) were also common. Most participants were overweight (median 30.0 kg/m^2^; IQR: 25.4 to 33.2). At baseline, 169 participants required only ambient oxygen or nasal cannula supplemental oxygen, and an additional 46 participants (18.7%) were requiring high flow nasal cannula (HFNC) or non-invasive positive pressure ventilation (NIPPV). (Table 1) Lastly, 40 participants (16.3%) required mechanical ventilation at the time of initial ultrasound scanning. During hospitalization, the most frequent treatments included dexamethasone (63.6% of participants), remdesivir (50.0%), or tocilizumab (9.1%).

**Table 1.** Participant baseline demographics.

| **Characteristic** | **Total (N=264)** |
| --- | --- |
| **Age —years, median (IQR)** | 58.56 (48.75, 68.00) |
| **Female — no. (%)** | 114 (43.18) |
| **Race — no. (%)** |  |
| Asian | 7 (2.65) |
| Black | 126 (47.73) |
| White | 80 (30.30) |
| Other | 49 (18.46) |
| **Ethnicity — no. (%)** |  |
| Hispanic | 44 (16.67) |
| Non-Hispanic | 220 (83.33) |
| **Smoking — no. (%)** |  |
| Never | 149 (56.44%) |
| Current | 23 (8.71%) |
| Former | 80 (30.30%) |
| **Median BMI — kg/m<sup>2</sup>, median (IQR)** | 30.00 (25.4, 33.2) |
| **Comorbidities — no. (%)** |  |
| Cancer | 25 (9.5) |
| Congestive heart failure | 87 (33.0) |
| COPD | 96 (36.4) |
| Diabetes | 112 (42.4) |
| Hypertension | 196 (74.2) |
| HIV/AIDS | 12 (4.6) |
| Liver Disease | 54 (20.5) |
| **Symptoms onset until LUS Scan — days, median (IQR)** | 9.29 (5.2, 14.3) |
| **Total lung zones scanned — median (IQR)** | 9 (7, 12) |
| **mLUS Score — median (IQR)** | 1.00 (0.50, 1.30) |
| **A-line lung fields — %, median (IQR)** | 0.75 (0.58, 0.92) |
| **B-line lung fields — %, median (IQR)** | 0.67 (0.38, 0.84) |
| **Pleural Line abnormality lung fields — %, median (IQR)** | 0.00 (0.00, 0.18) |

NIPPV: Non-invasive positive pressure ventilation; mLUS: mean lung ultrasound score

### Baseline cross-sectional differences in POCUS findings by severity

At enrollment, participants with severe illness were later in their course of illness (median: 15.79 days post symptom onset; IQR: 10.92 to 26.70 days) compared to those with moderately severe (median: 9.29 days; IQR: 7.03 to 13.92 days), or moderate illness (median 7.38 days; IQR 4.08 to 11.92 days) (Table 2). A lines were the most common finding among lung zones scanned (median 75.0% of lung fields; IQR, 58.3 to 91.7%), with a stepwise decrease in proportion of lung zones affected in moderately severe disease (median 69.7%; IQR, 51.8 to 87.1%) followed by severe disease at enrollment (54.6%; IQR, 25.0 to 66.7%) (Table 2). B lines were more likely to be present among those with severe disease (median 75.0%; IQR, 60.0 to 100%) or moderately severe disease (median 81.8%; IQR, 67.9 to 100%) compared to moderate cases (median 57.1%; IQR, 27.3 to 75.0%). Similarly, participants requiring mechanical ventilation at enrollment had higher percent of pleural line abnormalities (median 25.0%; IQR, 9.1 to 50) compared to moderately severe (median 0.0%; 0.0 to 15.6%) or moderate (median 0.0%; IQR 0.0 to 16.7%) disease. The mLUSS was lower for moderate disease (median 0.83; IQR, 0.33 to 0.80) compared to a stepwise increase in moderately severe disease (median 1.11; IQR, 1.00 to 1.50) followed by severe critical disease (1.25; IQR, 1.00 to 1.67). The Pearson correlation coefficient of the mLUSS between readers was high at 0.77 among 61 participants with a an available matched masked LUS read (Supplemental Figure S1).

**Table 2.**
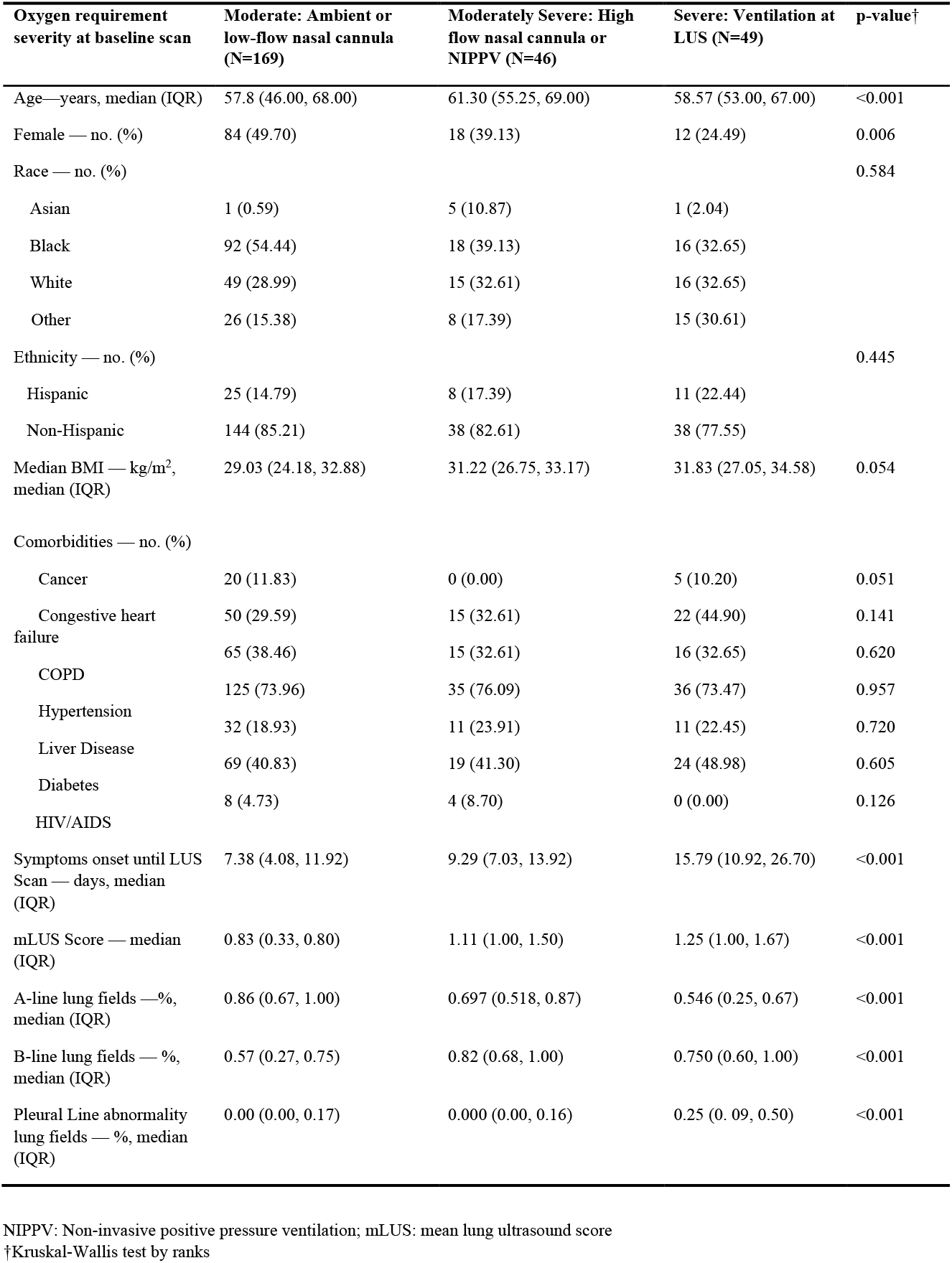
Cross-sectional differences in lung ultrasound findings by severity at baseline.

| Oxygen requirement severity at baseline scan | Moderate: Ambient or low-flow nasal cannula (N=169) | Moderately Severe: High flow nasal cannula or NIPPV (N=46) | Severe: Ventilation at LUS (N=49) | p-value† |
| --- | --- | --- | --- | --- |
| Age—years, median (IQR) | 57.8 (46.00, 68.00) | 61.30 (55.25, 69.00) | 58.57 (53.00, 67.00) | <0.001 |
| Female — no. (%) | 84 (49.70) | 18 (39.13) | 12 (24.49) | 0.006 |
| Race — no. (%) |  |  |  | 0.584 |
| Asian | 1 (0.59) | 5 (10.87) | 1 (2.04) |  |
| Black | 92 (54.44) | 18 (39.13) | 16 (32.65) |  |
| White | 49 (28.99) | 15 (32.61) | 16 (32.65) |  |
| Other | 26 (15.38) | 8 (17.39) | 15 (30.61) |  |
| Ethnicity — no. (%) |  |  |  | 0.445 |
| Hispanic | 25 (14.79) | 8 (17.39) | 11 (22.44) |  |
| Non-Hispanic | 144 (85.21) | 38 (82.61) | 38 (77.55) |  |
| Median BMI — kg/m<sup>2</sup>, median (IQR) | 29.03 (24.18, 32.88) | 31.22 (26.75, 33.17) | 31.83 (27.05, 34.58) | 0.054 |
| Comorbidities — no. (%) |  |  |  |  |
| Cancer | 20 (11.83) | 0 (0.00) | 5 (10.20) | 0.051 |
| Congestive heart failure | 50 (29.59) | 15 (32.61) | 22 (44.90) | 0.141 |
| COPD | 65 (38.46) | 15 (32.61) | 16 (32.65) | 0.620 |
| Hypertension | 125 (73.96) | 35 (76.09) | 36 (73.47) | 0.957 |
| Liver Disease | 32 (18.93) | 11 (23.91) | 11 (22.45) | 0.720 |
| Diabetes | 69 (40.83) | 19 (41.30) | 24 (48.98) | 0.605 |
| HIV/AIDS | 8 (4.73) | 4 (8.70) | 0 (0.00) | 0.126 |
| Symptoms onset until LUS Scan — days, median (IQR) | 7.38 (4.08, 11.92) | 9.29 (7.03, 13.92) | 15.79 (10.92, 26.70) | <0.001 |
| mLUS Score — median (IQR) | 0.83 (0.33, 0.80) | 1.11 (1.00, 1.50) | 1.25 (1.00, 1.67) | <0.001 |
| A-line lung fields —%, median (IQR) | 0.86 (0.67, 1.00) | 0.697 (0.518, 0.87) | 0.546 (0.25, 0.67) | <0.001 |
| B-line lung fields — %, median (IQR) | 0.57 (0.27, 0.75) | 0.82 (0.68, 1.00) | 0.750 (0.60, 1.00) | <0.001 |
| Pleural Line abnormality lung fields — %, median (IQR) | 0.00 (0.00, 0.17) | 0.000 (0.00, 0.16) | 0.25 (0.09, 0.50) | <0.001 |

NIPPV: Non-invasive positive pressure ventilation; mLUS: mean lung ultrasound score
†Kruskal-Wallis test by ranks

### Risk of disease progression

When evaluating the 28-day risk of progression to severe COVID-19, multiple baseline POCUS parameters were found to be associated with severity progression using Cox proportional hazards regression. Each point increase in the mLUSS was associated with disease progression to ICU-level care (aHR = 3.61; 95% CI: 1.27 to 10.22) and 28-day mortality (aHR = 3.10; 95% CI: 1.29 to 7.50), but not the composite outcome of ventilation or death (aHR = 2.45; 95% CI 0.81 to 11.02) (Figure 3 and Figure 4). Inference was unchanged when adjusting for total number of available lung zones with an increased risk of progression to ICU-level care (aHR = 3.80; 95% CI: 1.32 to 10.95) or death (aHR: 2.47; 95% CI: 1.10, 5.55), but not the composite outcome of ventilation or death (aHR: 2.96; 95% CI: 0.80 to 11.01). Similarly, inference was unchanged when excluding asymptomatic individuals with each increase in the mLUSS associated with risk of progression to ICU-level care (aHR = 3.07; 95% CI: 1.04 to 9.07), death (aHR: 2.94; 95% CI: 1.21 to 7.15), but not ventilation or death together (aHR: 2.69; 95% CI: 0.66 to 11.05). Lastly, when including days since symptom onset, there was an increased risk with each additional day (aHR: 1.007; 95% CI: 1.001 to 1.01), but the risk associated with each increase in mLUSS was similar (aHR: 3.86; 95% CI: 1.32 to 11.30) (Supplemental Table S1). There was no interaction observed between duration of symptoms and mLUSS (p=0.48) (data not shown).

**Figure 3.**
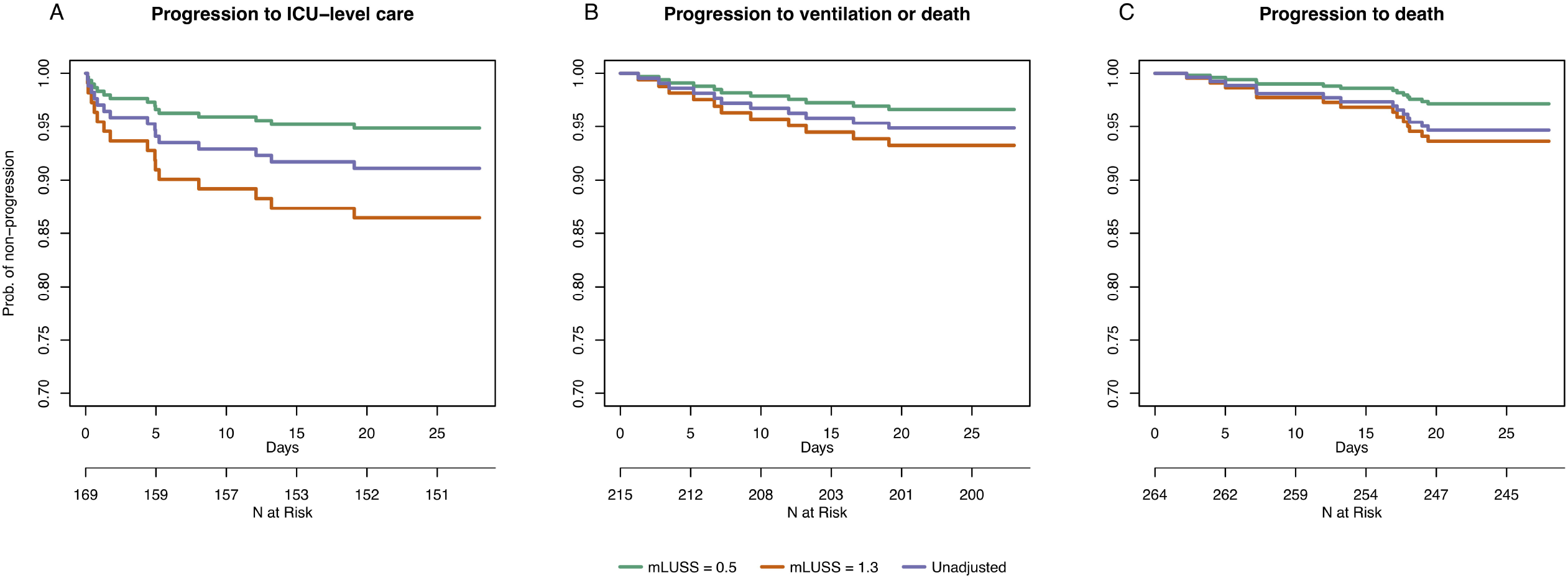
Kaplan-Meier time to progression by 75^th^ percentile (1.3) vs 25^th^ percentile (0.5) mLUSS scores from time of lung ultrasound scan.

**Figure 4.**
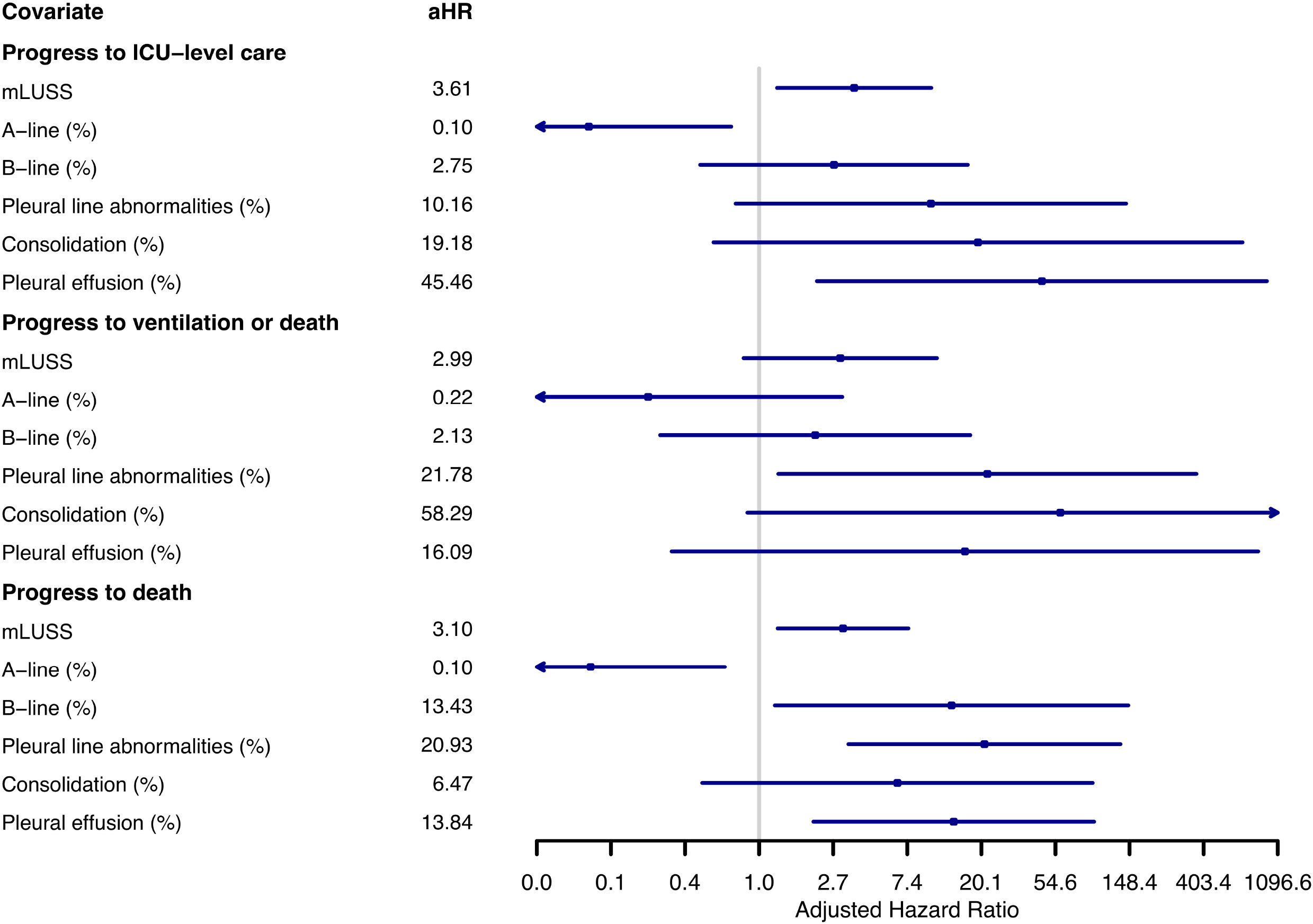
Forest Plot of hazard ratios of LUS parameters from individually fit models adjusting for age and sex for progression to severe disease.

Individual lung ultrasound characteristics were associated with disease progression. The presence of any type of B line was not associated with an increased risk of progression to ICU-level care (among those not on NIPPV, HFNC, or mechanical ventilation) (adjusted hazard ratio [aHR] = 2.75; 95% confidence interval [CI], 0.45 to 16.71) but was associated with 28-day mortality compared to those without any B lines (aHR = 13.43; 95% CI, 1.24 to 145.79) (Figure 4; Supplemental Table S1). Except for A lines, all studied individual POCUS parameters (i.e., B lines, pleural line changes, consolidations, and pleural effusions) had an increased risk of progression to ICU-level care, death, and death or ventilation but did not always meet statistical significance. Accordingly, A lines, which are generally present in the absence of B lines, were associated with a decreased risk of progression to ICU-level care (aHR 0.10; 95% CI, 0.02 to 0.69) and 28-day mortality (aHR = 0.10; 95% CI, 0.02-0.63). Pleural abnormalities were independently associated with mechanical ventilation plus death (aHR = 21.78; 95% CI, 1.30 to 365.95), and death (pleural line aHR = 20.93; 95% CI, 3.34 to 131.30).

## Discussion

We observed mLUSS and multiple individual LUS findings were associated with a subsequent increased oxygen requirement or death in a prospective cohort. Our results support LUS to identify hospitalized patients that may need a higher level of care or transport to centers with ICU beds available. While many studies to date have used retrospective clinical data, we conducted a prospective cohort study with standardized time points, probes, and protocols for image acquisition. The findings of this study support the use of lung ultrasound as a clinical tool that can improve triage by using individual POCUS abnormalities or mLUSS. Participants with more A lines on LUS are less likely to clinically deteriorate and participants with irregular pleural lines were more likely to have or to require higher levels of care. The mLUSS correlated well between ultrasound readers and could be used for patients with difficult to scan lung zones. These findings demonstrate the prognostic value of individual LUS findings or the mLUSS in assessing anticipated disease severity trajectories of COVID-19 without necessarily requiring all 12 lung zones.

Although there have been few large studies (5, 8), LUS has been shown to be associated with radiographic and clinical severity among adults hospitalized with COVID-19. Nouvenne et al. demonstrated B lines, pleural line irregularities and large parenchymal consolidations with correlated with CT findings and oxygen saturation.(14) In a systematic review of 43 studies the presence of focal, multifocal and/or confluent B lines and the presence of pleural irregularities were common among individuals with COVID-19.(15) Mechanistically, the degree and magnitude of LUS abnormalities throughout lung zones reflects the extent of lung disease and is intuitively directly related to severe disease trajectories. In one of the largest studies to date with matched CT scans, LUS compared to CT as a gold standard for severe COVD-19 had an area under the curve of 0.78 (CI 95% 0.68–0.87; p□<□0.001).(16) Rubio-Garcia and colleagues examined the LUS among 130 patients with COVID 19 and demonstrated an increased risk of mortality among individuals with a high modified LUS score (HR 5.25, 0.84–32.84) (9). The investigators however did not describe individual features of the LUS such as A lines, B lines and pleural disease and used a high cutoff to optimize sensitivity (9). While other studies have generally used a sum of all 12 lung zones (5, 8, 9, 17), our study found the risk estimates were unchanged when some lung fields were not obtainable due to clinical instability. Our study is consistent with prior publications and provides evidence that LUS can prognosticate hospitalized patients using available lung zones.

Adoption of LUS has varied in hospital settings largely a result of lack of familiarity as well as difference in approaches, techniques, and nomenclature (18). However, research personnel in our study were taught lung ultrasound using standardized protocols without prior experience. Ultrasound images were overread by multiple reviewers and correlated well. Most ultrasound operators were research coordinators who had no experience with ultrasound scanning prior to training for this study. This provided standardization of scans and reduced bias related to direct performance by medical caregivers. This suggests that LUS scanning could be expanded to non-clinicians, including but not limited to nursing staff, respiratory therapists, or medics in the field. For example, respiratory therapists or nursing staff could routinely perform LUS scans to obtain and document this valuable information, similar to lung auscultation performed at some centers. Alternatively, ultrasound technicians would be well-equipped to perform LUS scans with a standardized read by radiology. While the value of immediate information to a performing clinician should not be ignored or undervalued, extending the expertise of LUS performance to additional healthcare workers would be more scalable than LUS by clinicians alone.

Biomarker and therapeutic research has identified the importance of phase of disease as indicated by duration of symptoms (19). However, inference was unchanged after adjusting for duration of symptoms or interaction with duration in our Cox regression models. POCUS results appeared to be generalizable regardless of adjustment for days since symptom onset for determining risk of decompensation towards ICU-level care. Change in LUS findings were not evaluated here due to a limited sample size of repeat time events (data not shown), but studies are ongoing to evaluate longitudinal LUS for estimating risk of severe disease and treatment response.

There were limitations to the present study. First, not all participants were enrolled prior to admission, and as this was a hospital-based protocol, generally had a minimum requirement of oxygen. Patients were not always enrolled on the day of admission which may have diminished the effect size of differences in POCUS findings. Additionally, those hospitalized with incidental asymptomatic SARS-CoV-2 infection may be less comparable to those with moderate severity, but a sensitivity analysis was performed and inference about risk was unchanged. These factors led to sample size limitations in some of the survival analyses leading to wide confidence intervals, but the qualitative inference was consistent across outcomes and remains important. Further work is ongoing in ambulatory settings and additional sites with standardized follow-up to improve our understanding of the external validity and diagnostic accuracy among additional populations including non-hospitalized individuals with COVID 19. Lastly, while the mLUSS provide valuable prognostic information, additional lung ultrasound features such as consolidations or pleural line changes appear to be useful prognostic findings and should be evaluated for incorporation into models with subsequent validation. Future research with machine learning and unsupervised approaches can help optimize LUS for clinical use.

## Conclusion

Individual LUS findings and the mLUSS across available lung zones on lung POCUS are associated with ultimate oxygen requirement or death, independent of duration of illness among hospitalized patients.

## Data Availability

All data produced in the present study are available upon reasonable request to the authors.

## Acknowledgments

We thank the participants within the CCPSEI cohort study. Potential conflicts of interest: Y.C.M. receives research funding from Becton Dickinson, Quanterix, and Hologic and receives funding support to Johns Hopkins University from miDiagnostics.

## Conferences

This analysis has not been previously presented.

## Disclaimer

The contents of this article are the sole responsibility of the authors and do not necessarily reflect the views, assertions, opinions, or policies of the Henry M. Jackson Foundation for the Advancement of Military Medicine, Inc., the U.S. Department of Defense, the

## U.S. government, or any other government or agency

Mention of trade names, commercial products, or organizations does not imply endorsement by the U.S. government. The investigators have adhered to the policies for protection of human subjects as prescribed in 45 CFR 46.

