## Supplemental Table and Figure Legend for "Point-of-care lung ultrasound predicts severe disease and death due to COVID-19: a prospective cohort study"

**Supplemental Table S1.** Cox proportional hazard regression models for progression to ICU-level care adjusted for age and sex.

| | Dependent variable: Progression to ICU-level care | | | | | | |
|---|---|---|---|---|---|---|---|
| **Model** | (1) | (2) | (3) | (4) | (5) | (6) | (7) |
| **Covariates** | | | | | | | |
| mLUSS | 3.607** | | | | | | 3.858** |
| | (1.274, 10.217) | | | | | | (1.317, 11.302) |
| | p = 0.016 | | | | | | p = 0.014 |
| Age | 1.008 | 1.009 | 1.007 | 1.012 | 1.012 | 1.012 | 1.009 |
| | (0.971, 1.046) | (0.974, 1.045) | (0.973, 1.043) | (0.977, 1.047) | (0.978, 1.047) | (0.978, 1.048) | (0.971, 1.049) |
| | p = 0.677 | p = 0.629 | p = 0.682 | p = 0.513 | p = 0.493 | p = 0.492 | p = 0.654 |
| Male sex | 1.086 | 0.927 | 0.926 | 0.880 | 0.896 | 0.744 | 1.057 |
| | (0.382, 3.088) | (0.336, 2.557) | (0.332, 2.584) | (0.319, 2.431) | (0.324, 2.479) | (0.265, 2.089) | (0.369, 3.028) |
| | p = 0.878 | p = 0.884 | p = 0.884 | p = 0.806 | p = 0.833 | p = 0.576 | p = 0.918 |
| A-lines (% lung fields) | | 0.100** | | | | | |
| | | (0.015, 0.686) | | | | | |
| | | p = 0.020 | | | | | |
| B-lines (% lung fields) | | | 2.748 | | | | |
| | | | (0.452, 16.707) | | | | |
| | | | p = 0.273 | | | | |
| Pleural line abnormalities (% lung fields) | | | | 10.157* | | | |
| | | | | (0.728, 141.644) | | | |
| | | | | p = 0.085 | | | |
| Consolidation (% lung fields) | | | | | 19.181 | | |
| | | | | | (0.540, 680.960) | | |
| | | | | | p = 0.105 | | |

| | | |
|---|---|---|
| Pleural effusion (% lung fields) | 45.460**<br>(2.181, 947.701)<br>p = 0.014 | |
| Duration of symptoms (days) | | 1.0065**<br>(1.001, 1.012)<br>p = 0.030 |

Note: *p<0.1; **p<0.05; ***p<0.01

**Supplemental figure legend.**

**Supplemental Figure 1.** Correlation between two independent ultrasound clip readers for mLUSS (Pearson's $\rho$=0.77) .
