## Supplementary figures and images for "Point-of-care lung ultrasound predicts severe disease and death due to COVID-19: a prospective cohort study"

### Supplemental Figure S1.

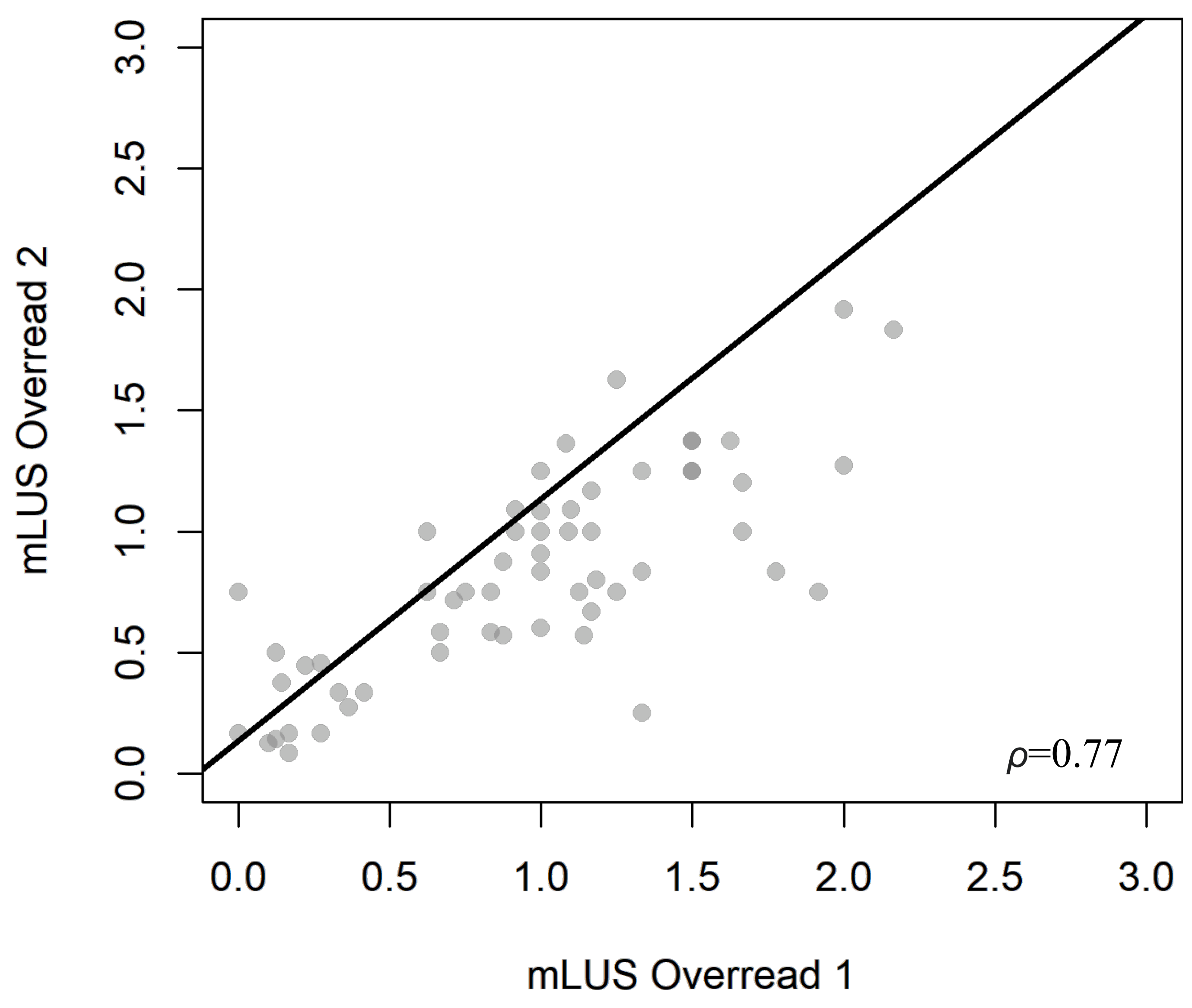
